# Immunomodulators and risk for breakthrough infection after third COVID-19 mRNA vaccine among patients with rheumatoid arthritis: A cohort study

**DOI:** 10.1101/2023.10.08.23296717

**Authors:** Abigail E. Schiff, Xiaosong Wang, Naomi J. Patel, Yumeko Kawano, Emily N. Kowalski, Claire E. Cook, Kathleen M.M. Vanni, Grace Qian, Katarina J. Bade, Alene A. Saavedra, Shruthi Srivatsan, Zachary K. Williams, Rathnam K. Venkat, Zachary S. Wallace, Jeffrey A. Sparks

**Affiliations:** Department of Medicine, Brigham and Women’s Hospital, Boston, MA, USA (60 Fenwood Road, Boston, MA, 02115); Division of Rheumatology, Inflammation, and Immunity, Brigham and Women’s Hospital, Boston, MA, USA (60 Fenwood Road, Boston, MA, 02115); Division of Rheumatology, Allergy, and Immunology, Massachusetts General Hospital, Boston, MA, USA (Rheumatology Associates, 55 Fruit Street, Boston, MA, 02114); Harvard Medical School, Boston, MA; Clinical Epidemiology Program, Mongan Institute, Department of Medicine, Massachusetts General Hospital, Boston, MA, USA (The Mongan Institute, 100 Cambridge Street, Suite 1600, Boston, MA, 02114); Tufts University School of Medicine, Boston, MA, USA (145 Harrison Ave, Boston, MA 02111)

**Author notes:** **<u>Corresponding author</u>,** Jeffrey A. Sparks MD MMSc, Division of Rheumatology, Inflammation, and Immunity Brigham and Women’s Hospital, 60 Fenwood Road, Suite 6016U Boston, MA 02115, @jeffsparks. Contributed equally (co-senior authors) (Zachary S. Wallace, Jeffrey A. Sparks).

**Keywords:** COVID-19, epidemiology, immunosuppression, reinfection

## Abstract

**Objectives:** To investigate COVID-19 breakthrough infection after third mRNA vaccine dose among patients with RA by immunomodulator drug class, and we hypothesized that CD20 inhibitors (CD20i) would have higher risk for breakthrough COVID-19 vs. TNF inhibitors (TNFi).

**Methods:** We performed a retrospective cohort study investigating breakthrough COVID-19 among RA patients at Mass General Brigham in Boston, MA, USA. Patients were followed from the date of 3rd vaccine dose until breakthrough COVID-19, death, or end of follow-up (18/Jan/2023). Covariates included demographics, lifestyle, comorbidities, and prior COVID-19. We used Cox proportional hazards models to estimate breakthrough COVID-19 risk by immunomodulator drug class. We used propensity score (PS) overlap-weighting to compare users of CD20i vs. TNFi.

**Results:** We analyzed 5781 patients with RA that received 3 mRNA vaccine doses (78.8% female, mean age 64.2 years). During mean follow-up of 12.8 months, 1173 (20.2%) had breakthrough COVID_19. Use of CD20i (adjusted HR 1.74, 95%CI 1.30-2.33) and glucocorticoid monotherapy (adjusted HR 1.47, 95%CI 1.09-1.98) were each associated with breakthrough COVID-19 compared to TNFi use. In the PS overlap-weighted analysis, CD20i users also had higher breakthrough COVID-19 risk than TNFi users (HR 1.62, 95%CI 1.02-2.56). A sensitivity analysis excluding patients with cancer or interstitial lung disease yielded similar findings.

**Conclusions:** We identified CD20i and glucocorticoid monotherapy as risk factors for breakthrough COVID-19 among patients with RA after a 3rd vaccine dose. This contemporary study highlights the real-world impact of blunted immune responses in these subgroups and the need for effective risk mitigation strategies.

**What is already known about this topic:**

- Patients with RA are at increased risk for COVID-19 breakthrough infection after two vaccine doses so a third dose is recommended to complete the initial series.
- Some immunomodulator medications, particularly CD20 inhibitors, can impact vaccine immunogenicity and waning.

**What this study adds:**

- CD20 inhibitor use was associated with increased risk of COVID-19 breakthrough infection in people with RA who received 3 vaccine doses compared to TNF inhibitor use.
- Glucocorticoid monotherapy was also associated with increased risk of COVID-19 breakthrough infection.

**How this study might affect research, practice or policy:**

- Patients with RA who are using CD20 inhibitors or glucocorticoid monotherapy should be prioritized for risk mitigation strategies after the initial vaccine series of 3 mRNA doses.
- The impact of additional vaccine doses, timing of medication dosing, and other protective measures will need further study.

## INTRODUCTION

In August 2021, the CDC recommended a third SARS-CoV-2 mRNA vaccine to complete the initial vaccine series for immunosuppressed patients who had previously received two mRNA doses^1^. Thus, many previous studies of breakthrough infection were performed using a 2-dose definition rather than the contemporary definition of 3 doses. Even after completing the initial vaccine series, some patients with rheumatoid arthritis (RA) remain at increased risk for breakthrough COVID-19 infection^2,3^ due to altered underlying immunity and use of immunomodulators which may lead to blunted immune responses to vaccination and infection^4^. Many immunomodulators, especially CD20 inhibitors (CD20i) and glucocorticoids, have been shown to decrease humoral responses to COVID-19 vaccines^4–8^, but may not affect T cell responses to the same extent^6,9^. As effective protection against infection can be difficult to predict from immune measurements alone, there is a need for well controlled studies to understand the contribution of individual immunomodulators to the risk of disease.

Previous studies of breakthrough COVID-19 in patients with systemic autoimmune diseases (SARDs) have some limitations. A retrospective cohort study^2^ investigated breakthrough infection and severity of infection after two mRNA doses, and found that patients with RA were at higher risk of breakthrough infection compared to patients without RA. Another study^10^ found that patients with rheumatic disease who had good vaccine responses by antibody titer after 2 vaccine doses had lower rates of breakthrough infections than inadequate or non-responders. <u>Boekel et al., Lancet Rheum 2022</u>^11^ found that patients on immunosuppressants had similar risk of breakthrough infection compared to those not on immunosuppressants, and found that seroconversion after vaccination was protective against breakthrough infection. Existing studies investigated small or heterogeneous cohorts^10,12–14^, did not have data by specific SARD diagnoses^15^, investigated lower numbers of COVID-19 vaccinations^2,3,16,17^, or did not compare immunosuppressant classes^2,3^.

Therefore, we investigated the risk of breakthrough COVID-19 among RA patients who had completed the initial series of 3 vaccine doses using disease-modifying antirheumatic drugs (DMARDs) or glucocorticoids. We hypothesized that users of CD20i would be at increased risk of breakthrough infection after 3 vaccine doses compared to users of TNFi.

## METHODS

### Study design and population

We performed a retrospective cohort study investigating immunomodulatory medications and risk of breakthrough infection after initial vaccine series (i.e., 3 mRNA vaccine doses) among patients with RA at Mass General Brigham (MGB). MGB is a multi-center healthcare system that includes a total of 14 hospitals, including two tertiary care hospitals (Massachusetts General Hospital and Brigham and Women’s Hospital), as well as other primary care and specialty outpatient centers in the greater Boston, Massachusetts area.

All patients included had RA, were taking at least one immunomodulatory medication, and had received at least three mRNA vaccine doses (Pfizer/BioNTech BNT162b2 or Moderna mRNA-1273) between 1/Feb/2021 and 6/Jul/2022. The end of follow-up was 18/Jan/2023. We restricted the analysis to residents of Massachusetts because of state laws mandating vaccine reporting. Recipients of the Janssen/Johnson and Johnson vaccine were excluded since this was infrequent and no longer recommended. The index date was the date of 3rd vaccine receipt. This study was approved by the MGB Institutional Review Board.

### Identification of patients with RA

We identified patients within MGB who were ≥18 years of age, were Massachusetts residents, had at least 2 ICD codes for RA within 2 years before initial COVID-19 vaccine dose, and received an immunomodulator medication before COVID-19 vaccine dose, as previously described^17^. The positive predictive value of this algorithm for RA is 90%. We queried the MGB centralized data warehouse, Research Patient Data Registry, to identify patients. We identified 5781 individuals who met these criteria (**Supplemental Figure 1**).

### Exposure variable: Immunomodulator use

We identified immunomodulator use as either 1) a prescription for a conventional synthetic, biologic, or targeted synthetic DMARD within 12 months of the index date or 2) a prescription for 30 pills of either oral prednisone or methylprednisolone within 6 months of the index date. For patients prescribed multiple immunomodulators, we used a hierarchy of medications to assign them to a single category (**Supplemental Table 1**). The hierarchy is as follows: CD20i, TNFi, IL-6 receptor inhibitor (IL-6Ri), cytotoxic T-lymphocyte associated protein 4 immunoglobulin (CTLA-4 Ig), Janus kinase inhibitors (JAKi), conventional synthetic DMARDs (csDMARDs) other than methotrexate or combination csDMARD, methotrexate monotherapy, antimalarial monotherapy, and glucocorticoid monotherapy. For example, a patient on a TNFi and methotrexate in combination would be classified in the TNFi group. The primary exposure of interest was that of CD20i use vs. TNFi use as the reference group. We chose this to limit possible confounding by indication since patients not on biologic or targeted synthetic DMARDs may be in an earlier phase of RA or have less severe disease, similar to a previous study^18^.

### Outcome variable: COVID-19 breakthrough infection

We identified patients with a positive test result for SARS-CoV-2 either by 1) a positive SARS-CoV-2 polymerase chain reaction (PCR) or antigen test from a nasopharyngeal or respiratory specimen at MGB, or 2) by other positive testing as noted by a COVID-19 flag 14 days or later after third vaccine dose. The MGB electronic health record (EHR) flags patients who reported positive testing either at home (such as patients who notified a clinician about a positive rapid antigen test through the secure patient portal) or outside of the MGB system (e.g., a patient with COVID-19 infection transferred from an outside hospital). As a secondary outcome, we identified patients who experienced severe outcomes, defined as hospitalization or death within 30 days of the initial positive test.

### Covariates

Baseline characteristics within 1 year of index date were extracted from the medical record using electronic query. We collected information on demographics (including age, sex, race/ethnicity as obtained from the electronic health record), smoking status, body mass index (BMI, in kg/m^2^), comorbidities as defined by international classification of diseases (ICD)-10 codes, previous COVID-19 infection, COVID-19 mRNA vaccine brands (all 3 doses Pfizer/BNT162b2, all 3 doses Moderna/mRNA-1273, or mix), median zip code area-level household income, number of encounters with the healthcare system, number of hospitalizations, and RA duration in years based on initial ICD-10 code in the EHR. The Charlson Comorbidity Index (CCI)^19^ was calculated using all available data from comorbidities as surveyed by ICD-10 codes in the one year prior to the index date^17^. We also included calendar time. We divided this into periods based on circulating variants: pre-Omicron (1/Feb/2021 to 16/Dec/2021), initial Omicron wave (17/Dec/2021 to 31/Jan/2022), and additional Omicron wave (1/Feb/2022 to 18/Jan/2023). We used an indicator variable for those with missing race. We used multiple imputation for those missing BMI^20^. There were no other missing data.

### Statistical analysis

We presented descriptive statistics of the overall study sample and dichotomized those who did and did not experience COVID-19 breakthrough infection after the index date of 3rd mRNA vaccine dose. Categorical variables were presented as frequency (percentage), and continuous variables were presented as mean (standard deviation) or median (interquartile range), as appropriate. We reported the frequency and proportion of people who experienced COVID-19 breakthrough infection, hospitalization, and death among the users of each category of immunomodulator drug class. For descriptive purposes, we also reported post-baseline variables, such as tixagevimab/cilgavimab (pre-exposure prophylaxis) and additional vaccine doses. We did not adjust for these post-baseline variables since they may have been mediators, rather than confounders, that were on the causal pathway between immunomodulator medication use and breakthrough infection risk.

We assessed the associations of immunomodulator drug class for RA with the risk of breakthrough infection using unadjusted and multivariable Cox regression models to estimate hazard ratios (HR) and 95% confidence intervals (CI). Follow-up time accrued from the date of 3rd vaccine dose. Patients were censored at COVID-19 breakthrough infection (primary outcome), death, or end of study (18/Jan/2023), whichever came first. We calculated the incidence rates and 95%CIs for each stratum of the immunomodulator medication variable. The primary multivariable model adjusted for age, sex, race, Hispanic, BMI, smoking status, CCI, interstitial lung disease (ILD), previous vaccination, previous COVID-19, calendar time, median area-level household income, previous encounters, previous hospitalizations, and RA duration. We chose these factors based on our *a priori* understanding of the associations of these factors with exposures and/or outcomes. The additional multivariable model also adjusted for concomitant oral glucocorticoids; thus, glucocorticoid monotherapy users were excluded. We plotted cumulative probability curves for COVID-19 breakthrough infection.

We also performed propensity score (PS)-overlap weighted analysis for the primary comparison of CD20i vs. TNFi use to further address potential confounding by indication. We included all covariates at the index date and calculated the PS using logistic regression models and adopted an overlap weighting approach to balance baseline characteristics^21,22^. To account for temporal changes based on vaccine availability and variants, we included continuous calendar time of the index date during the study in the model. We displayed characteristics before and after PS overlap-weighting using descriptive statistics and compared CD20i vs TNFi groups by standardized mean difference. We then calculated mean follow-up time, incidence rates, risk difference, HR, and associated 95%CIs for COVID-19 breakthrough infection, comparing CD20i and TNFi groups. We plotted the cumulative incidence curve of breakthrough COVID-19 for these two groups.

Since CD20 inhibitors can be used for other indications like cancer and interstitial lung disease (ILD) that can also increase risk of COVID-19 infection^23,24^, we performed a sensitivity analysis in the PS overlap-weighted analysis that excluded participants with cancer and ILD.

We tested the proportional hazard assumption in all analyses using Schoenfeld residuals. We considered a two-sided p value <0.05 as statistically significant in all analyses. All analyses were performed using SAS v9.4.

## RESULTS

### Study sample and baseline characteristics

We included 5781 patients with RA that received at least 3 mRNA vaccine doses (**Table 1, Supplemental Figure 1**). A total of 1173 (20.3%) had COVID-19 after the index date (date of third vaccination). A total of 4311 (74.6%) patients used csDMARDs, 2470 (42.7%) used biologic DMARDs [including 1727 (29.9%) on TNFi and 172 (3.0%) on CD20i], 490 (8.5%) used JAKi, and 792 (13.7%) used oral glucocorticoids.

**Table 1.** Baseline demographic and clinical characteristics at date of 3^rd^ mRNA vaccine receipt among patients with rheumatoid arthritis (n=5781).

| Characteristic | All RA patients who received at least 3 mRNA vaccine doses (n=5781) | COVID-19 after index date (n=1173) | No COVID-19 after index date (n=4608) |
| --- | --- | --- | --- |
| Mean age, years (SD) | 64.2 (14.2) | 62.3 (14.9) | 64.6 (14.0) |
| Female sex | 4555 (78.8%) | 933 (79.5%) | 3622 (78.6%) |
| Race |  |  |  |
| Asian | 169 (2.9%) | 28 (2.4%) | 141 (3.1%) |
| Black or African American | 268 (4.6%) | 43 (3.7%) | 225 (4.9%) |
| Other or multiple | 294 (5.1%) | 50 (4.3%) | 244 (5.3%) |
| Unknown | 181 (3.1%) | 36 (3.1%) | 145 (3.2%) |
| White | 4869 (84.2%) | 1016 (86.6%) | 3853 (83.6%) |
| Hispanic or Latinx ethnicity | 334 (5.8%) | 53 (4.5%) | 281 (6.1%) |
| Median Charlson Comorbidity Index (median [IQR]) | 1 (1, 3) | 2 (1, 4) | 1 (1, 3) |
| Comorbidities |  |  |  |
| Hypertension | 2586 (44.7%) | 593 (50.6%) | 1993 (43.3%) |
| Diabetes | 828 (14.3%) | 176 (15.0%) | 652 (14.2%) |
| Malignancy excluding non-melanoma skin cancer | 759 (13.1%) | 190 (16.2%) | 569 (12.4%) |
| Asthma | 786 (13.6%) | 231 (19.7%) | 555 (12.0%) |
| Coronary artery disease | 662 (11.5%) | 161 (13.7%) | 501 (10.9%) |
| Interstitial lung disease | 303 (5.2%) | 74 (6.3%) | 229 (5.0%) |
| Median RA duration, years (IQR) | 5 (2, 10) | 5 (2, 10) | 5 (2, 10) |
| Immunomodulatory medications |  |  |  |
| Conventional synthetic DMARDs | 4311 (74.6%) | 862 (73.5%) | 3449 (74.9%) |
| Methotrexate | 2669 (46.2%) | 543 (46.3%) | 2126 (46.1%) |
| Antimalarials | 1686 (29.2%) | 342 (29.2%) | 1344 (29.2%) |
| Hydroxychloroquine monotherapy | 803 (13.9%) | 158 (13.5%) | 645 (14.0%) |
| Leflunomide | 429 (7.4%) | 74 (6.3%) | 355 (7.7%) |
| Sulfasalazine | 308 (5.3%) | 62 (5.3%) | 246 (5.3%) |
| Mycophenolate mofetil/mycophenolic acid | 78 (1.4%) | 20 (1.7%) | 58 (1.3%) |
| Azathioprine | 58 (1.0%) | 15 (1.3%) | 43 (0.9%) |
| Cyclophosphamide | 9 (0.2%) | 1 (0.1%) | 8 (0.2%) |
| Biologic DMARDs | 2470 (42.7%) | 537 (45.8%) | 1933 (42.0%) |
| TNF inhibitor | 1727 (29.9%) | 364 (31.0%) | 1363 (29.6%) |
| CTLA-4 immunoglobulin | 282 (4.9%) | 51 (4.4%) | 231 (5.0%) |
| IL-6 receptor inhibitor | 238 (4.1%) | 51 (4.4%) | 187 (4.1%) |
| CD20 inhibitor | 172 (3.0%) | 55 (4.7%) | 117 (2.5%) |
| JAK inhibitor | 490 (8.5%) | 99 (8.4%) | 391 (8.5%) |
| Oral glucocorticoid | 792 (13.7%) | 205 (17.5%) | 587 (12.7%) |
| Previous COVID-19 | 340 (5.9%) | 30 (2.6%) | 310 (6.7%) |
| Calendar time |  |  |  |
| Feb 1, 2021 – Dec 16, 2021 | 4348 (75.2%) | 945 (80.6%) | 3403 (73.8%) |
| Dec 17, 2021 – Jan 31, 2022 | 1115 (19.3%) | 193 (16.5%) | 922 (20.0%) |
| Feb 1, 2022 – Jul 6, 2022 | 318 (5.5%) | 35 (3.0%) | 283 (6.1%) |
| Median zip-code area annual household income (\$USD, IQR) | 88,395 (68,865,110,025) | 92,327 (73,525,113,417) | 87,516 (67,589,107,444) |
| Median number of healthcare encounters in previous year (IQR) | 14 (1, 31) | 23 (6, 41) | 12 (1, 29) |
| Hospitalization in previous year | 483 (8.4%) | 120 (10.2%) | 363 (7.9%) |

CD20, cluster of differentiation 20; CTLA-4, cytotoxic T lymphocyte-associated 4; DMARDs, disease-modifying antirheumatic drugs; IL-6, interleukin-6; IQR, interquartile index; JAK, janus kinase; RA, rheumatoid arthritis; SD, standard deviation; TNF, tumor necrosis factor; USD, United States dollars

Mean age was 64.2 years (SD 14.2) among all patients, with a mean of 62.3 years (SD 14.9) for those with breakthrough COVID-19 and 64.6 years (SD 14.0) for those without. Of the patients who had breakthrough COVID, 86.6% were White, while 83.6% of patients without were White. The percent of patients with ILD was 6.3% in those with breakthrough COVID and 5.0% without. Median RA duration was similar at 5 (IQR 2,10) years in both groups. Median number of healthcare encounters in the previous year was 23 (IQR 6, 41) in those with breakthrough COVID and 12 (IQR 1, 29) in those without. Approximately three-fourths of patients (75.2%) received their third vaccine dose before December 16, 2021, when the Omicron wave started. Patients with vs. without COVID-19 after the index date had a higher CCI [median 2 (IQR 1,4) vs. 1 (IQR 1,3)] and higher rates of hypertension (50.6% vs. 43.3%), malignancy (16.2% vs. 12.4%), asthma (19.7% vs 12.0%), and coronary artery disease (13.7% vs. 10.9%). Patients who had COVID-19 after the third vaccine were less likely to have had previous COVID-19 infection (2.6% vs. 6.7%).

### COVID-19 outcomes

The overall frequencies and proportions experiencing COVID-19 breakthrough infection, hospitalizations, and death were 1173 (20.3%), 62 (1.1%), and 8 (0.1%), respectively (**Table 2**). CD20i users (n=172) more frequently had COVID-19 and severe outcomes compared to users of other DMARD categories: 55 (32.0%) cases of COVID-19, 4 (2.3%) hospitalizations or deaths, and 1 (0.6%) death. Users of glucocorticoid monotherapy (n=173) also had higher proportions of COVID-19 and severe outcomes compared to other DMARD categories; 49 (28.3%) with COVID-19, 6 (3.5%) with hospitalization or death, and 1 (0.6%) death. Conversely, TNFi users (n=1651) had 341 (20.7%) cases of COVID-19, 13 (0.8%) hospitalization or death, and 3 (0.2%) deaths. **Supplemental Table 2** shows post-baseline variables, including the numbers of patients who received tixagevimab/cilgavimab, and 4th, 5th, and 6th vaccine doses by baseline immunomodulator group.

**Table 2.** SARS-CoV-2 infections and outcomes by baseline immunomodulator group.

|  | **COVID-19** | **Hospitalization** | **Death** |
| --- | --- | --- | --- |
| Overall (n=5781) | 1173 (20.3%) | 62 (1.1%) | 8 (0.1%) |
| Glucocorticoid monotherapy (n=173) | 49 (28.3%) | 5 (2.8%) | 1 (0.6%) |
| Antimalarial monotherapy (n=807) | 161 (20.0%) | 11 (1.3%) | 1 (0.1%) |
| Methotrexate monotherapy (n=1500) | 298 (19.9%) | 18 (1.2%) | 1 (0.1%) |
| Other csDMARD or combination csDMARD (n=532) | 86 (16.2%) | 8 (1.5%) | 1 (0.2%) |
| TNF inhibitor (n=1651) | 341 (20.7%) | 10 (0.6%) | 3 (0.2%) |
| CTLA-4 Ig (n=260) | 45 (17.3%) | 2 (0.8%) | 0 (0.0%) |
| JAK inhibitor (n=451) | 87 (19.3%) | 5 (1.1%) | 0 (0.0%) |
| IL-6 receptor inhibitor (n=235) | 51 (21.7%) | 0 (0.0%) | 0 (0.0%) |
| CD20 inhibitor (n=172) | 55 (32.0%) | 3 (1.7%) | 1 (0.6%) |

CD20, cluster of differentiation 20; csDMARD, conventional synthetic disease modifying antirheumatic drug; CTLA-4 Ig, cytotoxic T lymphocyte-associated 4 immunoglobulin; IL-6, interleukin-6; JAK, janus kinase; SARS-CoV-2, severe acute respiratory syndrome coronavirus 2; TNF, tumor necrosis factor

### Hazard ratios for COVID-19 by immunomodulator class

**Table 3** includes the incidence rates and HRs for COVID-19 by immunomodulator class. The incidence rate (95% CI) per 1000 person-months for COVID-19 infection for users of glucocorticoid monotherapy was 24.4 (17.6, 31.2) with an unadjusted HR (95% CI) of 1.47 (1.09, 1.98), for other csDMARD or combination csDMARD the IR was 12.6 (9.9, 15.3) with an HR was 0.78 (0.62, 0.99), and for CD20i the IR was 27.7 (20.4, 35.0) with an HR of 1.74 (1.30, 2.33), all of which were statistically significant compared to TNFi users. Other immunomodulator classes had similar rates of COVID-19 infection after the third mRNA vaccine dose compared to TNFi, including antimalarial monotherapy, methotrexate monotherapy, CTLA-4 Ig, JAKi, and IL-6Ri. These differences persisted for users of CD20i and glucocorticoid monotherapy in the primary multivariable models when compared to TNFi users (HR 1.65 [1.22, 2.24] and HR 1.53 [1.09, 2.14], respectively). The findings regarding the association of CD20i use with breakthrough infection persisted when adjusting for glucocorticoid use (HR of 1.64 [1.21, 2.23]). Cumulative incidence of COVID-19 by immunomodulator class is presented in **Figure 1**, with the highest cumulative incidence for CD20i and glucocorticoid monotherapy.

**Figure 1.**
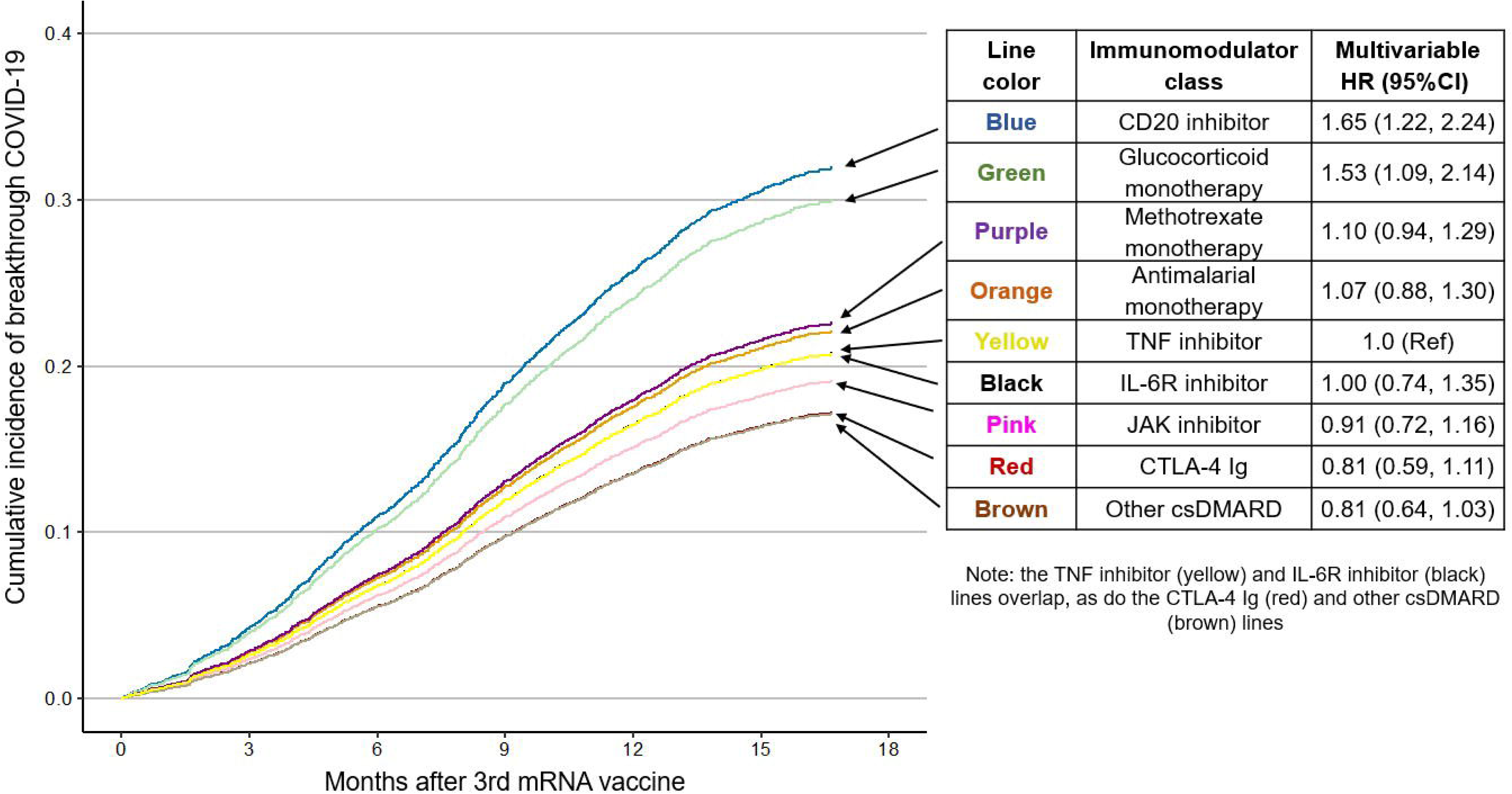
Cumulative incidence and hazard ratio for breakthrough COVID-19 by immunomodulator use at 3rd COVID mRNA vaccine dose. Adjusted for age, sex, race, Hispanic ethnicity, body mass index, smoking status, Charlson Comorbidity Index, rheumatoid arthritis duration, interstitial lung disease, vaccine type (Pfizer, Moderna, mix), previous COVID-19, calendar time, median zip-code area level household income, number of healthcare encounters in previous years, and number of hospitalizations in previous year.

**Table 3.** Hazard ratios for COVID-19 by immunomodulator use at 3^rd^ COVID mRNA vaccine dose among rheumatoid arthritis patients (n=5781).

| Immunomodulator class | COVID-19 cases | Person-months | Incidence rate (95%CI) per 1000 person-months | Unadjusted HR (95%CI) | Multivariable model 1* HR (95%CI) | Multivariable model 2** HR (95%CI) |
| --- | --- | --- | --- | --- | --- | --- |
| Glucocorticoid monotherapy (n=173) | 49 | 2,010.0 | 24.4 (17.6, 31.2) | **1.47 (1.09, 1.98)** | **1.53 (1.09, 2.14)** | N/A |
| Antimalarial monotherapy (n=807) | 161 | 9,984 | 16.1 (13.6, 18.6) | 1.00 (0.83, 1.20) | 1.07 (0.88, 1.30) | 1.07 (0.89, 1.30) |
| Methotrexate monotherapy (n=1500) | 298 | 19,443.5 | 15.3 (13.6, 17.1) | 0.97 (0.83, 1.13) | 1.10 (0.94, 1.29) | 1.10 (0.94, 1.29) |
| Other csDMARD or combination csDMARD (n=532) | 86 | 6,830.0 | 12.6 (9.9, 15.3) | **0.78 (0.62, 0.99)** | 0.81 (0.64, 1.03) | 0.80 (0.63, 1.02) |
| TNF inhibitor (n=1651) | 341 | 21,655.5 | 15.8 (14.1, 17.4) | 1.0 (Ref) | 1.0 (Ref) | 1.0 (Ref) |
| CTLA-4 Ig (n=260) | 45 | 3,473.4 | 13.0 (9.2, 16.7) | 0.82 (0.60, 1.12) | 0.81 (0.59, 1.11) | 0.81 (0.59, 1.10) |
| JAK inhibitor (n=451) | 87 | 5,996.9 | 14.5 (11.5, 17.6) | 0.92 (0.73, 1.17) | 0.91 (0.72, 1.16) | 0.91 (0.72, 1.16) |
| IL-6R inhibitor (n=235) | 51 | 3,115.4 | 16.4 (11.9, 20.9) | 1.04 (0.78, 1.39) | 1.00 (0.74, 1.35) | 1.00 (0.75, 1.34) |
| CD20 inhibitor (n=172) | 55 | 1,986.0 | 27.7 (20.4, 35.0) | **1.74 (1.30, 2.33)** | **1.65 (1.22, 2.24)** | **1.64 (1.21, 2.23)** |

Bolded values are statistically significant.
\*Adjusted for age, sex, race, Hispanic, BMI, smoking status, CCI, ILD, previous vaccination, Previous COVID-19, calendar time, median area-level household income, previous encounters, previous hospitalizations, and RA duration
\*\*Additionally adjusted for oral glucocorticoids
BMI, body mass index; CCI, Charlson comorbidity index; CD20, cluster of differentiation 20; CI, confidence interval; csDMARD, conventional synthetic disease modifying antirheumatic drug; CTLA-4 Ig, cytotoxic T lymphocyte-associated 4 immunoglobulin; HR, hazard ratio; IL-6R, interleukin 6 receptor; ILD, interstitial lung disease; RA, rheumatoid arthritis; Ref, reference; TNF, tumor necrosis factor

### Propensity score overlap weighted analysis for CD20 inhibitor vs. TNF inhibitor

**Table 4** shows the distribution of the baseline characteristics of the CD20i and TNFi users before and after PS overlap-weighting. The incidence rate of COVID-19 infection per 1000 person-months (95% CI) in the CD20i group was 27.3 (19.3, 35.4), compared to 16.6 (10.7, 22.5) in the TNFi group, with a risk difference of 10.75 (0.75, 20.74). In the propensity-score weighted analysis, the findings from the propensity-score adjusted analysis were confirmed. CD20i users had a hazard ratio for breakthrough COVID-19 of 1.64 (1.03, 2.59) compared to TNFi users (**Figure 2, Supplemental Table 3**).

**Figure 2.**
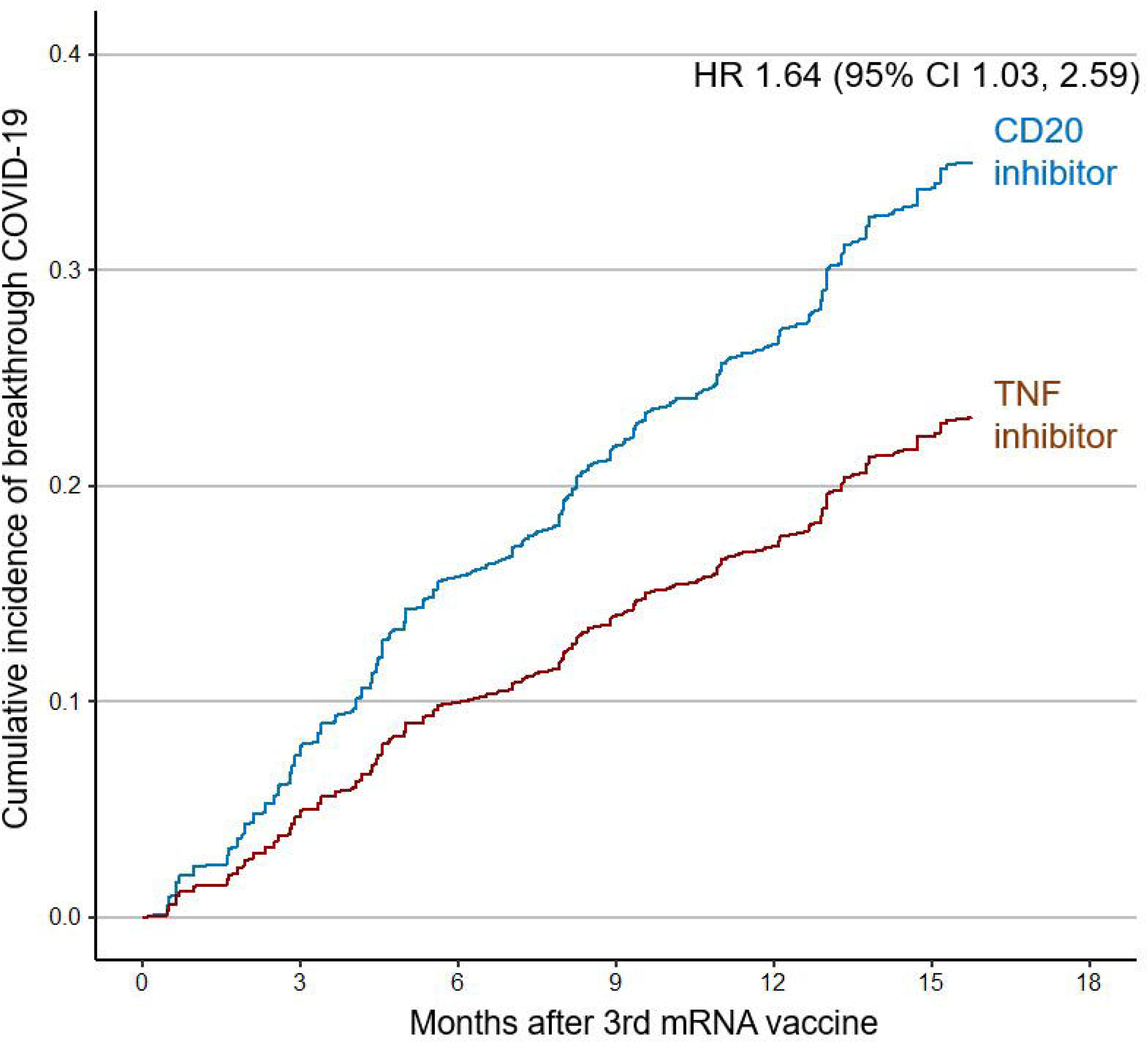
Propensity-score weighted cumulative incidence of COVID-19 after 3rd mRNA vaccine, comparing CD20 inhibitor users vs. TNF inhibitor users. Propensity score weighted analysis includes age, sex, race, Hispanic ethnicity, BMI, smoking status, Charlson Comorbidity Index, interstitial lung disease, malignancy, rheumatoid arthritis duration, concomitant csDMARDs, concomitant glucocorticoid use, vaccine type (Pfizer, Moderna, mix), previous COVID-19, calendar time, median zip-code area level household income, number of healthcare encounters in previous year, and number of hospitalizations in previous year.

**Table 4.** Baseline characteristics according to CD20 inhibitor and TNF inhibitor use among RA patients at 3^rd^ mRNA vaccine dose, before and after propensity score weighting.

|  | Before PS overlap weighting |  |  | After PS overlap weighting |  |  |
| --- | --- | --- | --- | --- | --- | --- |
|  | CD20i | TNFi | Standardized difference | CD20i | TNFi | Standardized difference |
| n | 172 | 1651 | N/A | 172 | 1651 | N/A |
| Mean age, years (SD) | 63.77 (13.52) | 60.63 (14.30) | 0.23 | 62.97 (12.32) | 62.97 (4.22) | <0.001 |
| Female | 80.8% | 77.2% | 0.088 | 81.5% | 81.5% | <0.001 |
| Race |  |  |  |  |  |  |
| White | 76.2% | 84.5% | 0.21 | 77.6% | 77.6% | <0.001 |
| Black or African American | 8.1% | 4.2% | 0.16 | 7.3% | 7.3% | <0.001 |
| Asian | 2.9% | 3.0% | 0.0072 | 2.8% | 2.8% | <0.001 |
| Other or multiple | 9.9% | 5.0% | 0.19 | 9.2% | 9.2% | <0.001 |
| Unknown | 2.9% | 3.3% | 0.021 | 3.1% | 3.1% | <0.001 |
| Hispanic or Latinx ethnicity | 6.4% | 5.8% | 0.027 | 6.8% | 6.8% | <0.001 |
| Body mass index (mean, SD, kg/m<sup>2</sup>) | 27.68 (6.17) | 28.53 (6.85) | 0.13 | 27.68 (5.58) | 27.68 (1.90) | <0.001 |
| Smoking status |  |  |  |  |  |  |
| Never | 69.8% | 74.0% | 0.093 | 70.7% | 70.7% | <0.001 |
| Past | 25.6% | 22.4% | 0.074 | 24.4% | 24.4% | <0.001 |
| Current | 4.7% | 3.6% | 0.051 | 4.9% | 4.9% | <0.001 |
| Charlson Comorbidity Index (median [IQR]) | 2 (1, 6) | 1 (1, 3) | 0.44 | 2 (1, 5) | 2 (1, 5) | <0.001 |
| Interstitial lung disease | 23.3% | 3.4% | 0.61 | 16.5% | 16.5% | <0.001 |
| Malignancy excluding non-melanoma skin cancer | 19.8% | 8.1% | 0.34 | 17.0% | 17.0% | <0.001 |
| Concomitant conventional synthetic DMARDs | 47.1% | 57.0% | 0.20 | 49.0% | 49.0% | <0.001 |
| Concomitant oral glucocorticoids | 15.1% | 8.7% | 0.20 | 14.1% | 14.1% | <0.001 |
| Previous COVID-19 immunity before index date |  |  |  |  |  |  |
| Previous COVID-19 | 5.8% | 5.0% | 0.038 | 6.1% | 6.1% | <0.001 |
| Previous vaccine types |  |  |  |  |  |  |
| Pfizer for all first 3 doses | 48.8% | 50.1% | 0.025 | 49.5% | 49.5% | <0.001 |
| Moderna for all first 3 doses | 46.5% | 43.7% | 0.056 | 45.6% | 45.6% | <0.001 |
| Mix of Pfizer or Moderna | 4.7% | 6.2% | 0.068 | 4.9% | 4.9% | <0.001 |
| Median Calendar time (Day, IQR) | 245.5 (197, 289.5) | 246 (198, 288) | 0.021 | 245 (197, 294) | 249 (198, 289) | <0.001 |
| Median zip-code area household income (\$USD, IQR) | 88,155.5 (65,412, 105,996.5) | 89,452 (70,163, 110,025) | 0.094 | 88,640 (66,681, 106,332) | 88,395 (66,091, 106,332) | <0.001 |
| Median number of healthcare encounters in previous year (IQR) | 22 (3, 41) | 14 (4, 29) | 0.32 | 22 (3, 40) | 19 (6, 39) | <0.001 |
| Number of hospitalizations in previous year (mean, SD) | 0.42 (1.72) | 0.18 (0.92) | 0.17 | 0.36 (1.45) | 0.36 (0.39) | <0.001 |
| Median RA duration (years, IQR) | 8 (2, 12) | 6 (2, 11) | 0.12 | 8 (3, 12) | 7 (2, 12) | <0.001 |

CD20i, cluster of differentiation 20 inhibitors; CTLA-4, cytotoxic T lymphocyte-associated 4; DMARDs, disease-modifying antirheumatic drugs; IL-6, interleukin-6; IQR, interquartile index; JAK, janus kinase; kg, kilograms; PS, propensity score; RA, rheumatoid arthritis; SD, standard deviation; TNFi, tumor necrosis factor inhibitors; USD, United States dollars

In a sensitivity analysis excluding patients with ILD or cancer from the propensity score-weight analysis, the association between CD20i use vs TNFi use with breakthrough infection remained similar (HR 1.62 [0.92, 2.87]).

## DISCUSSION

In this large cohort study of patients with RA, we found that CD20i and glucocorticoid monotherapy use were associated with higher risk of breakthrough COVID-19 compared to TNFi after the initial vaccine series of three mRNA doses. These novel findings take into account the current definition of breakthrough infection after the initial vaccine series, which includes 3 doses in this vulnerable immunosuppressed population^25^. Further, we include data collected during the Omicron wave of the COVID-19 pandemic, highlighting the relevance of our findings to the contemporary circulating variants. Our findings emphasize the importance of prioritizing people with RA using CD20i for risk mitigation strategies, such as additional vaccinations, behavior modification, and preventive therapies.

Patients with RA have an increased risk of COVID infection, hospitalization, and death compared to those without RA^26^, even after vaccination^3^. The most widely used CD20i, rituximab, has been shown to be associated with worse COVID-19 outcomes in RA patients in a large cohort that did not examine breakthrough infections^27^, and was associated with decreased rates of positive antibodies after three COVID vaccines^28^. The third dose of a COVID vaccine has been shown to reduce overall rates of breakthrough infection in the general population^29^ and in immunocompromised individuals without analysis of immunomodulator class^30^. Key studies that looked at breakthrough infection have used a definition of two vaccine doses ^2,17,31–33^. However, to our knowledge we are the first group to have investigated breakthrough infection risk by immunomodulator class after 3 vaccine doses in patients with RA. Our study shows that users of CD20i and glucocorticoid monotherapy have an increased risk of COVID-19 even after 3 vaccine doses.

Decreased immune protection from breakthrough infection is likely related to impaired immune responses to COVID-19 and other vaccinations observed in CD20i users^4,34^. Rituximab use in RA patients decreases humoral responses to influenza vaccination^35^, and these responses are worse when the vaccine was given 6 months after administration of rituximab than when given 6 days before the medication^36^. In another study of RA patients given rituximab, B cell mediated responses (pneumococcal polysaccharide and the neoantigen KLH), but not T cell mediated responses (tetanus toxoid or delayed type hypersensitivity responses to Candida albicans), were decreased by rituximab use^37^. The impact of glucocorticoid use on vaccine immunogenicity is less well understood^34^. For example, a study of glucocorticoid use and influenza vaccination found no impact on antibody titers^38^.

In addition to their associations with risk of breakthrough COVID-19 infection, immunomodulator use is also associated with worse acute outcomes of COVID-19. Both CD20i and glucocorticoid monotherapy use greater than an equivalent of prednisone 10 mg daily have been associated with higher rates of mortality from COVID-19 in people with systemic autoimmune disease^39^. It is possible they may also lead to greater risks of post-acute sequelae of COVID (PASC)^40^. Some individuals with immunosuppression exhibit prolonged infection, which can cause morbidity and mortality for the patient, as well as prolonged shedding and accelerated viral evolution, which may contribute to the ongoing immune evasion of SARS-CoV-2 and pose a public health threat^41–43^.

Our study has several strengths. We constructed a large contemporary cohort of patients with RA using immunomodulators linked to vaccine administration records from Massachusetts and comprehensive data on covariates. We restricted our analysis to patients with RA to limit confounding by indication of medications and used propensity score-based methods to confirm our findings. Unlike previous literature, we focused on participants who have been infected with COVID-19 at least 2 weeks after they received their third vaccine dose, which meets the current US CDC definition of breakthrough infection. There are also limitations to the study. First, we used a rule-based approach to identify RA patients so there may be some misclassification. However, the positive predictive value of this approach is 90%^17^, and it would not be feasible to perform medical record review to confirm RA diagnosis for all patients for a cohort of this size. Second, administrative data in the EHR was used to identify immunomodulator medications used by individual participants. It is possible that the participants may not have been adherent or may have changed therapies or that their therapies may have been held or adjusted around the time of vaccine dose. Misclassification of the use of immunomodulators would likely bias towards the null and not explain the findings. Third, glucocorticoid dose was not available to investigate so we cannot determine if there is a threshold of glucocorticoid dose associated with breakthrough infection risk. Fourth, we may not have identified all COVID-19 infections, especially when participants used home rapid antigen tests that may not have been entered into the medical record, when the infection was mild, or when the patient was not aware of the infection. This could lead to an underestimate of the incidence rate. We find it unlikely that this would vary by immunomodulator use group so should not explain positive findings. Fifth, we did not have information on RA severity but were able to adjust for RA duration and concomitant csDMARD and glucocorticoid use. Finally, this study was conducted in a single hospital system in the Northeast US and the majority of patients were non-Hispanic White. Further studies are needed in more diverse populations and in different geographic areas.

In this study evaluating the impact of immunomodulator use for RA on the risk of COVID-19 that included the Omicron phase of the pandemic, we identified CD20i and glucocorticoid monotherapy use as risk factors for breakthrough COVID-19. This contemporary study highlights the excess risk associated with glucocorticoid monotherapy or CD20i use for RA treatment and identifies important subgroups for counseling and additional risk mitigation strategies.

## FUNDING SOURCE AND COMPETING INTERESTS

### Contributors

AES, ZSW, and JAS designed the study, were responsible for acquisition, analysis and interpretation of data, and drafted and revised the article. XW was involved in the analysis and interpretation of the data. NJP, YK, ENK, CEC, KMMV, GQ, KJB, AS, SS, ZKW, and RV were involved in the data acquisition, interpretation, and revision of the manuscript. JAS and ZSW are joint senior authors. All authors approved the final version of the article. JAS accepts full responsibility for the work and the conduct of the study, had access to the data, and controlled the decision to publish.

### Funding/Support

NJP is supported by the Rheumatology Research Foundation. ZSW is funded by NIH/NIAMS (K23 AR073334 and R03 AR078938). JAS is funded by the National Institute of Arthritis and Musculoskeletal and Skin Diseases (grant numbers R01 AR080659, R01 AR077607, P30 AR070253, and P30 AR072577), the R. Bruce and Joan M. Mickey Research Scholar Fund, and the Llura Gund Award funded by the Gordon and Llura Gund Foundation. The funders had no role in the decision to publish or preparation of this manuscript. The content is solely the responsibility of the authors and does not necessarily represent the official views of Harvard University, its affiliated academic health care centers, or the National Institutes of Health.

### Competing Interests

NJP reports consulting fees from FVC Health unrelated to this work. ZSW reports research support from Bristol-Myers Squibb and Principia/Sanofi and consulting fees from Viela Bio, Zenas BioPharma, and MedPace. JAS has received research support from Bristol Myers Squibb and performed consultancy for AbbVie, Amgen, Boehringer Ingelheim, Bristol Myers Squibb, Gilead, Inova Diagnostics, Janssen, Optum, Pfizer, and ReCor unrelated to this work. All other authors report no competing interests.

### Patient and public involvement

Patients and/or the public were not involved in the design, or conduct, or reporting, or dissemination plans of this research.

### Patient consent for publication

Not applicable.

### Ethics approval

This study was approved by the Mass General Brigham Institutional Review Board (2020P000833).

### Data availability statement

Data are available on reasonable request. This study includes patient data from Mass General Brigham. The data that support the findings of this study may be made available upon reasonable request by contacting the corresponding author, JAS.

## Notes

### Author Declarations

Mass General Brigham Institutional Review Board of Mass General Brigham gave ethical approval for this work (2020P000833).

