## Supplemental Table for "Immunomodulators and risk for breakthrough infection after third COVID-19 mRNA vaccine among patients with rheumatoid arthritis: A cohort study"

*Contributed equally (co-senior authors)

**Affiliations**

Boston, MA, 02114)

^6^ Tufts University School of Medicine, Boston, MA, USA (145 Harrison Ave, Boston, MA 02111)

**Supplemental material**

**Supplemental Table 1.** Immunomodulator classes presented in hierarchical order.

**Supplemental Figure 1.** Identification of RA patients who had received at least 3 SARS-CoV-2 vaccinations.

**Supplemental Table 2.** Post-baseline variables, overall and by immunomodulator class.

**Supplemental Table 3**. Propensity score weighted analysis comparing CD20 inhibitor users vs. TNF inhibitor users for COVID-19 risk among RA patients.

**Supplemental Table 1.** Immunomodulator classes presented in hierarchical order.

CD20 inhibitor

IL-6 receptor inhibitor

JAK inhibitor

CTLA-4 immunoglobulin

TNF inhibitor

Conventional synthetic DMARDs other than methotrexate or combination csDMARDs

Methotrexate monotherapy

Antimalarial monotherapy

Glucocorticoid monotherapy

**Supplemental Figure 1.** Identification of RA patients who had received at least 3 SARS-CoV-2 vaccinations.

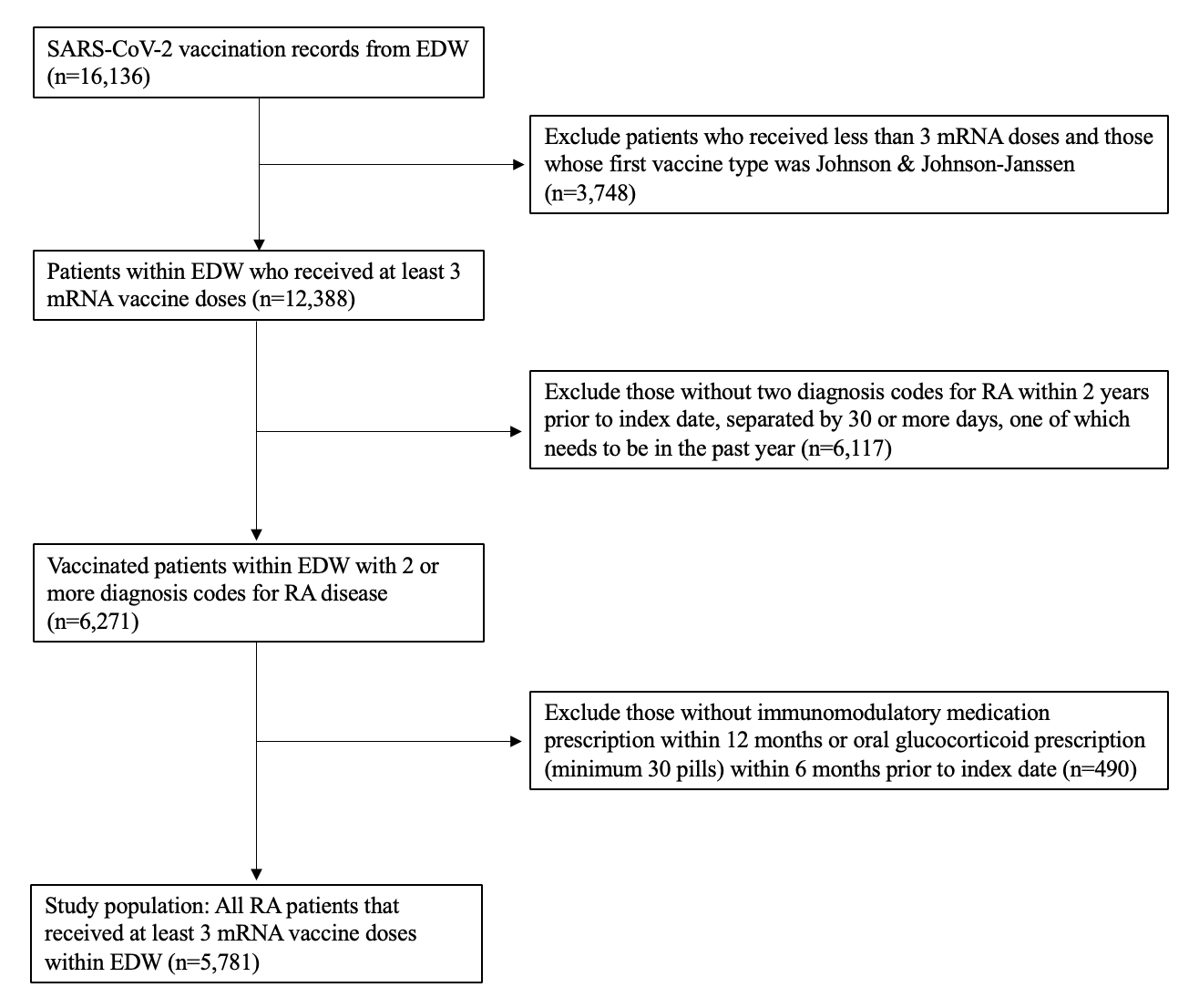

**Supplemental Table 2.** Post-baseline variables, overall and by immunomodulator class.

|  | **Received tixagevimab/cilgavimab** | **Received 4th vaccine dose** | **Received 5th vaccine dose** | **Received 6th vaccine dose** |
| --- | --- | --- | --- | --- |
| Overall (n=5781) | 119 (2.1%) | 1731 (30.0%) | 125 (2.2%) | 6 (0.1%) |
| Glucocorticoid monotherapy (n=173) | 0 (0.0%) | 39 (22.5%) | 0 (0.0%) | 0 (0.0%) |
| Antimalarial monotherapy (n=807) | 4 (0.5%) | 199 (24.7%) | 6 (0.7%) | 0 (0.0%) |
| Methotrexate monotherapy (n=1500) | 10 (0.7%) | 444 (30.0%) | 26 (1.7%) | 0 (0.0%) |
| Other csDMARD or combination csDMARD (n=532) | 4 (0.8%) | 150 (28.2%) | 6 (1.1%) | 0 (0.0%) |
| TNF inhibitor (n=1651) | 31 (1.9%) | 535 (32.4%) | 55 (3.3%) | 3 (0.2%) |
| CTLA-4 Ig (n=260) | 11 (4.2%) | 94 (36.2%) | 9 (3.5%) | 1 (0.4%) |
| JAK inhibitor (n=451) | 6 (1.3%) | 139 (30.8%) | 5 (1.1%) | 0 (0.0%) |
| IL-6R inhibitor (n=235) | 5 (2.1%) | 81 (34.5%) | 13 (5.5%) | 1 (0.4%) |
| CD20 inhibitor (n=172) | 48 (27.9%) | 50 (29.1%) | 5 (2.9%) | 1 (0.6%) |

Row percents are displayed.

CD20, cluster of differentiation 20; csDMARD, conventional synthetic disease modifying antirheumatic drug; CTLA-4 Ig, cytotoxic T lymphocyte-associated 4 immunoglobulin; IL-6R, interleukin-6 receptor; JAK, janus kinase; TNF, tumor necrosis factor

**Supplemental Table 3**. Propensity score weighted analysis comparing CD20 inhibitor users vs. TNF inhibitor users for COVID-19 risk among RA patients.

|  | **COVID-19 cases** | | **Mean follow-up (months)** | | **IR per 1000 person-months (95%CI)** | | **Risk difference (95%CI) person-months** | **HR (95%CI) CD20i vs. TNFi** |
| --- | --- | --- | --- | --- | --- | --- | --- | --- |
|  | **CD20i** | **TNFi** | **CD20i** | **TNFi** | **CD20i** | **TNFi** |  |  |
| n | 172 | 1651 | 172 | 1651 | 172 | 1651 |  |  |
| Primary analysis | 55 | 341 | 12 | 13 | 27.3 (19.3, 35.4) | 16.6 (10.7, 22.5) | **10.75 (0.75, 20.74)** | **1.64 (1.03, 2.59)** |
| Sensitivity analysis |  |  |  |  |  |  |  |  |
| Excluded cancer and ILD | 31 | 294 | 12 | 13 | 25.2 (15.9, 34.6) | 15.4 (8.6, 22.2) | 9.86 (-1.67, 21.39) | 1.62 (0.92, 2.87) |

Bolded values are statistically significant.

*Propensity score includes the following: age, sex, race, Hispanic, BMI, smoking status, CCI, ILD, cancer, csDMARDs, oral glucocorticoids, previous vaccination, Previous COVID-19, calendar time, median zip-code area level household income, number of encounters, number of hospitalizations, and RA duration

BMI, body mass index; CCI, Charlson comorbidity index; CD20i, cluster of differentiation 20 inhibitor; CI, confidence interval; csDMARD, conventional synthetic disease modifying antirheumatic drug; HR, hazard ratio; ILD, interstitial lung disease; IR, incidence rate; RA, rheumatoid arthritis; TNFi, tumor necrosis factor inhibitor
